# Number Needed to Vaccinate with a Novel Tuberculosis Vaccine to Prevent Tuberculosis in High-Risk Populations, United States

**DOI:** 10.64898/2026.05.11.26352950

**Authors:** Jessica E. Rothman, Kenneth G. Castro, Benjamin A. Lopman, Neel R. Gandhi, Kristin N. Nelson

**Author notes:** **Address for correspondence:** Jessica E. Rothman, Department of Epidemiology, Rollins School of Public Health, Emory University, 1518 Clifton Rd NE, Atlanta, GA 30322, USA.

## Abstract

We estimated the number needed to vaccinate (NNV) with an M72/AS01_E_-like vaccine to prevent one tuberculosis case in U.S. high-risk groups. Targeted vaccination of *Mycobacterium tuberculosis*–infected persons yielded NNVs of 217 (persons with HIV) to 2,486 (U.S.-born), within the range of established adult vaccines.

**Summary line:** Targeted vaccination of *Mycobacterium tuberculosis*-infected high-risk persons with an M72/AS01_E_-like vaccine could prevent one tuberculosis case per 217 (HIV) to 2,486 (U.S.-born) persons vaccinated, within the range of established adult vaccines.

## Introduction

U.S. tuberculosis (TB) incidence has increased each year since 2021, reversing decades of decline (*1*). TB burden is concentrated among non-U.S.-born persons, who account for about 75% of cases, and among persons with diabetes, HIV, or end-stage renal disease (ESRD), who together account for approximately 40% of cases (*2,3*). About 85% of U.S. TB cases are attributed to reactivation of longstanding *Mycobacterium tuberculosis (Mtb*) infection rather than recent transmission (*3,4*). While treatment of *Mtb* infection with preventive drug therapy (often referred to as tuberculosis preventive treatment, or TPT) can reduce progression risk, uptake of this intervention remains suboptimal due to adherence challenges and medication side effects (*5*). Vaccines that prevent progression to TB disease may offer an alternative approach to reduce TB incidence in high-risk populations.

The M72/AS01_E_ candidate, currently in Phase III trials, demonstrated approximately 50% efficacy in preventing progression to TB disease among *Mtb*-infected adolescents and adults in high-burden settings (*6*). Whether such a vaccine could meaningfully contribute to TB control in low-burden settings such as the United States has not been established.

We estimated NNV with an M72/AS01_E_-like vaccine to prevent one TB case among U.S. high-risk populations under two vaccination scenarios: all-comers and targeted.

### The Study

We estimated the NNV to prevent one case of TB in the U.S. by nativity (U.S.-born and non-U.S.-born) and by high-risk medical conditions (diabetes, HIV, and ESRD). From the National Tuberculosis Surveillance System (NTSS), we obtained TB case counts stratified by risk group and the proportion attributed to reactivation, as determined by CDC’s plausible source case method (*7*). We adapted previously published back-calculation methodology (*8*) to estimate the number of individuals with *Mtb* infection within each risk group, dividing reactivation-attributed cases by published group-specific reactivation rates (*3*). We evaluated NNV under two scenarios: (1) all-comers vaccination of all individuals within each high-risk group regardless of *Mtb* infection status, and (2) targeted vaccination of only those with confirmed *Mtb* infection. NNV under all-comers vaccination was calculated as the population size divided by the product of TB cases and vaccine efficacy. Under targeted vaccination, the numerator was the estimated *Mtb*-infected subpopulation. We assumed 50% vaccine efficacy based on the M72/AS01_E_ Phase IIb trial (*6*) and a 3-year time horizon matching trial follow-up. We constructed 95% CIs by parametric bootstrapping (1,000,000 iterations). Equations and full distributional assumptions are provided in the Technical Appendix. In sensitivity analyses, we repeated calculations at vaccine efficacy of 40%, 60%, 70%, and 80%. We conducted analyses in R version 4.5.2.

Under targeted vaccination, NNV was lowest for the high-risk medical-condition groups (Table). For people living with HIV (PLWH) and *Mtb* infection, 217 individuals would need to be vaccinated to prevent one TB case (95% CI 95–755). Targeted NNV was 369 (95% CI 133–1,370) for persons with ESRD and 1,991 (95% CI 960–6,800) for persons with diabetes. Among non-U.S.-born persons with *Mtb* infection, targeted NNV was 2,158 (95% CI 1,200–7,100), similar to the U.S.-born estimate of 2,486 (95% CI 660–10,100). For the total U.S. population with *Mtb* infection, targeted NNV was 2,240 (95% CI 1,100–7,600).

Under all-comers vaccination, NNVs were substantially higher across all groups (Table). For the total U.S. population, 70,350 persons would need to be vaccinated to prevent one TB case (95% CI 41,500–229,000). All-comers NNV was approximately 19-fold lower for non-U.S.-born persons (13,185; 95% CI 7,800–42,700) than for U.S.-born persons (252,021; 95% CI 148,500–824,000). Among medical-condition groups, all-comers NNV was lowest for persons with HIV (5,429; 95% CI 3,200–17,800), followed by ESRD (6,361; 95% CI 3,700–20,800) and diabetes (28,365; 95% CI 16,700–92,200).

Across vaccine-efficacy assumptions of 40%–80%, NNV scaled inversely with efficacy (Appendix Table 2). Under the most conservative assumption (VE = 40%), targeted NNVs ranged from 269 for persons with HIV to 3,088 for U.S.-born persons. Targeted-vaccination NNVs fell within the range of NNVs reported for established U.S. adult vaccines, including pneumococcal, HPV, and influenza vaccination (Figure; Appendix Table 3).

## Conclusions

We estimated NNV for an M72/AS01_E_-like vaccine to prevent TB in U.S. high-risk groups. Targeted vaccination of persons with *Mtb* infection yielded NNVs 6-to 101-fold lower than all-comers vaccination, ranging from 217 for persons with HIV to 2,486 for U.S.-born persons. These values fell within the range of NNVs reported for established U.S. adult vaccines such as PCV-13 (234–1,620), HPV vaccination against cervical cancer (324), and influenza vaccination (*9–11*). The only other published TB-vaccine NNV estimates, derived from dynamic transmission modeling of age-targeted vaccination in China, ranged from 230 to 1,022 (*12*). Despite differences in setting, methodology, and target population, these estimates also fall within the range of accepted vaccines. Although no universal NNV threshold defines the utility of a vaccination program (*13*), these comparisons place TB vaccination within the range of efficiency seen for existing programs.

Our analysis has several limitations. NNV captures direct vaccine effects only, but because *Mtb* transmission in the United States is rare, indirect effects are unlikely to substantially alter these estimates. We assumed the M72/AS01_E_ Phase IIb vaccine efficacy estimate (*6*) would apply to the U.S. populations evaluated here, though efficacy may be lower among immunocompromised groups such as persons with HIV or ESRD, and the lower-VE sensitivity scenarios may be most relevant for these subgroups. Our NNV estimates assume a 3-year time horizon matching Phase IIb trial follow-up. If vaccine-induced protection wanes beyond this period, long-term NNVs would be higher. Comparator NNVs for other vaccines reflect universal or age-targeted programs, limiting direct comparison with our risk-group targeted estimates.

The lower NNV under targeted vaccination suggests that programs incorporating *Mtb* infection screening (e.g., interferon-gamma release assay testing) in high-risk groups could substantially improve the efficiency of TB vaccination, although this approach must be balanced against screening cost and feasibility (*14*). Future work should evaluate the costeffectiveness of vaccination strategies relative to TPT and update these estimates as Phase III efficacy data become available.

## Supporting information

Technical Appendix

## Data Availability

Analysis code is available at https://github.com/jrothman27/tb-nnv-us. Aggregate TB case counts are publicly available in CDC's annual Reported Tuberculosis in the United States reports; individual-level NTSS data are available from CDC by request.

## Acknowledgments

This study was funded by an American Lung Association Catalyst Award (CA-943920), an NIH K01 (NIH/NIAID K01AI166093-01A1), and a developmental grant from the NIH Center for AIDS Research at Emory University (P30 AI050409) to K.N.N. The work was also supported by an NIH K24 (K24 AI114444) to N.R.G. and the Emory/Georgia Tuberculosis Research Advancement Center (P30 AI168386). J.E.R. was supported by the Ferguson Fellowship. The funders had no role in study design, conduct, analysis, or presentation of results.

---

**Table.**
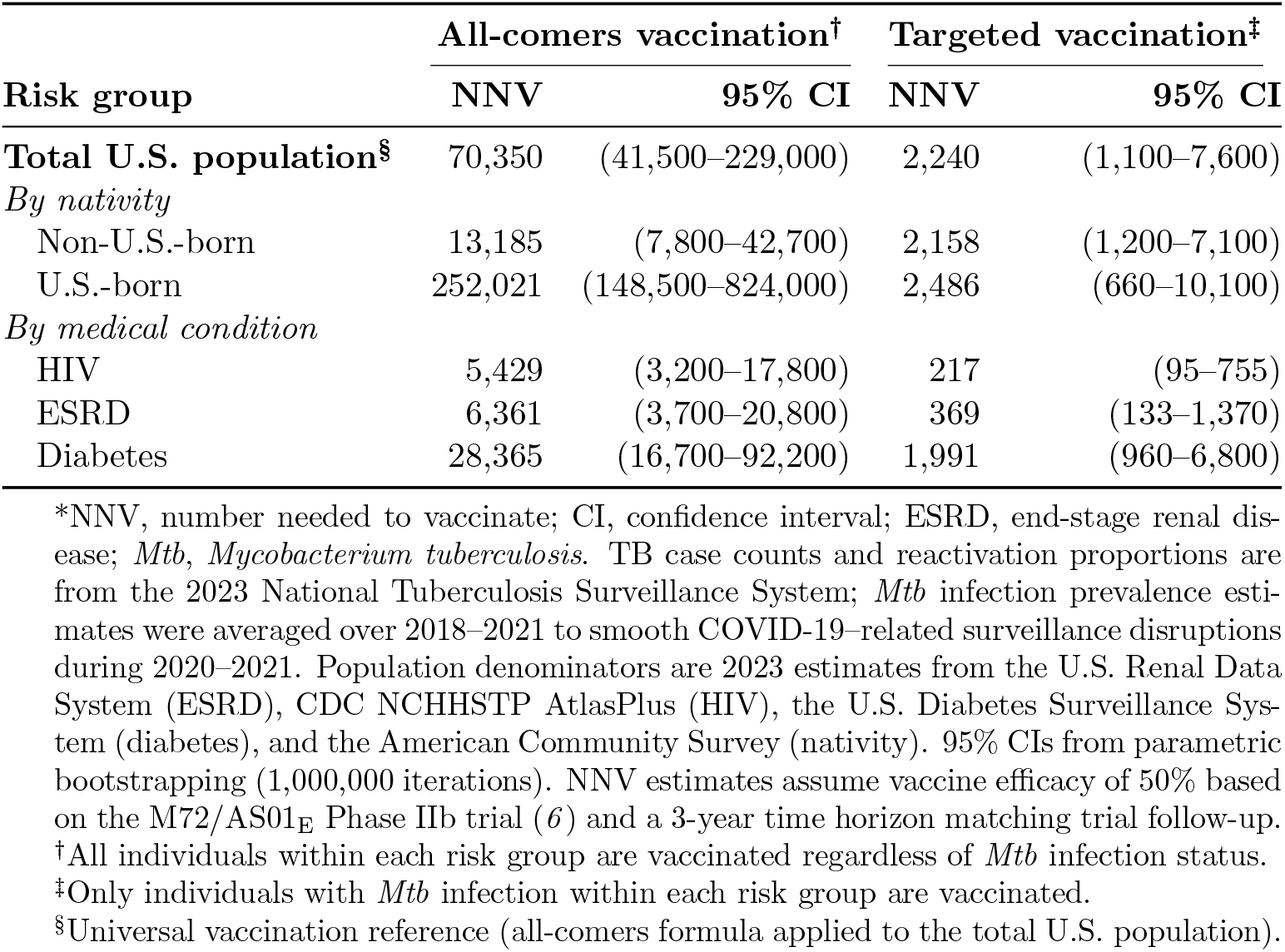
Number needed to vaccinate to prevent one tuberculosis case by risk group at 50% vaccine efficacy in U.S. populations*.

**Figure.**
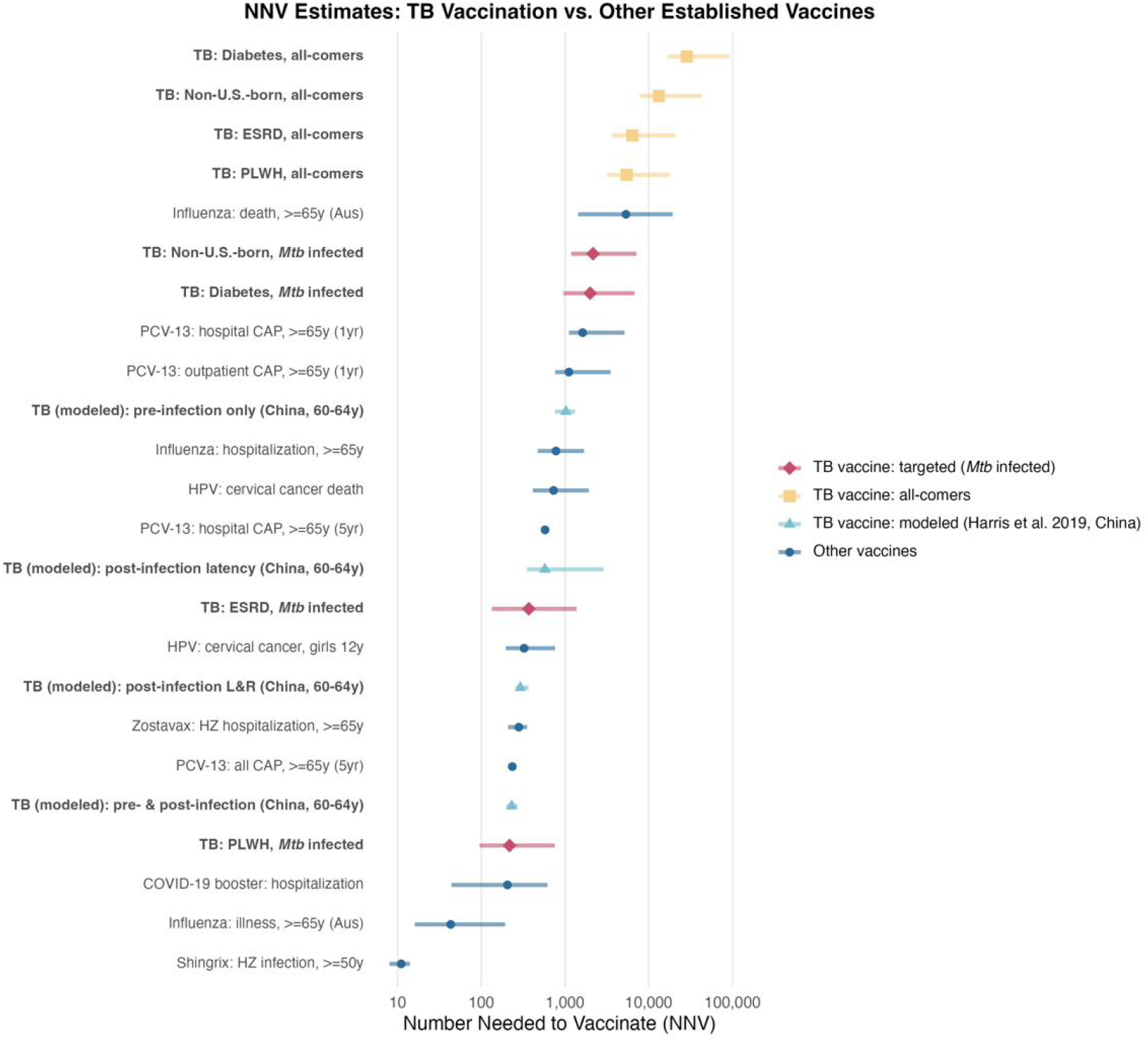
Number needed to vaccinate (NNV) estimates for an M72/AS01_E_-like tuberculosis (TB) vaccine in U.S. high-risk populations, compared with NNVs for established adult vaccines. TB-vaccine NNVs are shown for both targeted vaccination (vaccinating only persons with *Mtb* infection) and all-comers vaccination (vaccinating all individuals in a risk group regardless of infection status). Modeled TB-vaccine estimates from Harris et al. (*12*) reflect dynamic transmission modeling of age-targeted vaccination in China and are shown for context only. NNV is plotted on a log scale; horizontal lines indicate 95% confidence intervals. ESRD, end-stage renal disease; PLWH, persons living with HIV; HZ, herpes zoster; CAP, community-acquired pneumonia; HPV, human papillomavirus; PCV-13, 13-valent pneumococcal conjugate vaccine; L&R, latent and recovered.

