## Supplementary material for "Number Needed to Vaccinate with a Novel Tuberculosis Vaccine to Prevent Tuberculosis in High-Risk Populations, United States": Technical Appendix

### Data Sources

**Tuberculosis cases.** We obtained 2023 TB case counts and the proportion of cases that can be attributed to reactivation (reactivation proportions) for U.S. high-risk groups from the National Tuberculosis Surveillance System (NTSS). *Mtb* infection prevalence estimates were averaged over 2018–2021 to adjust for COVID-19–related disruptions in TB surveillance during 2020–2021.

**Reactivation rates.** We used risk group-specific reactivation rates (rate of progression from *Mtb* infection to TB disease) and corresponding 95% confidence intervals (CIs) from 2011–2012 NHANES infection data measuring interferon-gamma release assay (IGRA) positivity (*1*).

**Population denominators.** Population size estimates in 2023 were obtained from the NIDDK US Renal Data System (USRDS) for ESRD (*2*), NCHHSTP AtlasPlus for HIV (*3*), US Diabetes Surveillance System for diabetes (*4*), and the American Community Survey (ACS) for nativity (*5*) (Appendix Table 1).

**Vaccine efficacy.** For this analysis, we used vaccine efficacy estimates from the M72/AS01$_{\mathrm{E}}$ Phase IIb clinical trial, which demonstrated 49.7% efficacy (95% CI 2.1%–74.2%) in preventing progression to TB at 3 years post-vaccination among adolescents and adults in Kenya, South Africa, and Zambia with *Mtb* infection confirmed by IGRA positivity (*6*). We rounded this to 50% vaccine efficacy (VE) for our analysis.

### Estimating *Mtb* Infection Burden

We adapted previously published back-calculation methodology (*7*,*8*) to estimate the number of individuals with *Mtb* infection within each risk group. This approach divides the number of TB cases attributed to reactivation by the corresponding reactivation rate.

After initial *Mtb* infection, individuals may progress to TB through two pathways: recent transmission (progression within 2 years of infection) or reactivation (progression after years of latent infection). The CDC uses the plausible source case method, which combines genotypic and epidemiological data to classify TB cases as resulting from either recent transmission or reactivation (*9*). From NTSS, we obtained TB case data stratified by risk group and the proportion of TB cases attributed to reactivation, as determined by the plausible source case method (*9*,*10*). We then applied previously published risk-group-specific reactivation rates (*1*) to calculate the number of *Mtb* infections in each group, $i$:

$$M_i = \frac{\text{Reactivation TB cases}_i}{\text{Reactivation rate}_i} \tag{1}$$

### NNV Calculations

We calculated NNV under the two vaccination scenarios described in the main text, with the universal estimate serving as a reference. For all-comers vaccination (vaccinating all individuals within each risk group regardless of *Mtb* infection status):

$$\text{NNV}_i^{\text{all-comers}} = \frac{N_i}{t_i \times v} \tag{2}$$

For targeted vaccination (vaccinating only those with confirmed *Mtb* infection within each risk group):

$$\text{NNV}_i^{\text{targeted}} = \frac{M_i}{t_i \times v} \tag{3}$$

In Equations 1–3, $i$ denotes the specific risk group, $t_i$ is the number of TB cases, $N_i$ is the population size, $M_i$ is the number of *Mtb* infections estimated via Equation 1, and $v$ is the vaccine efficacy estimate.

#### Bootstrap Confidence Intervals

We constructed 95% confidence intervals for the NNV estimates using parametric bootstrapping (1,000,000 iterations). For all-comers vaccination, population size was treated as fixed, with uncertainty in NNV derived from the number of TB cases (modeled using a Poisson distribution) and vaccine efficacy (modeled using a Beta distribution parameterized from the M72/AS01$_E$ trial confidence interval). For targeted vaccination, the estimated number of *Mtb* infections was additionally treated as a random variable (lognormal distribution) to account for uncertainty in the back-calculation.

**Appendix Table 1.** Input parameters by risk group for NNV calculations.

| Risk Group | Population Size (2023) | TB Cases (2023) | *Mtb* Infections (2018–21 avg) | *Mtb* Prev. (%) | TB Incidence (per 100k) |
|---|---|---|---|---|---|
| **Total U.S. population** | 336,806,231 | 9,633 | 10,722,811 | 3.18 | 2.9 |
| **By nativity** | | | | | |
| Non-U.S.-born | 47,831,411 | 7,299 | 7,828,550 | 16.37 | 15.3 |
| U.S.-born | 287,083,485 | 2,292 | 2,831,426 | 0.99 | 0.8 |
| **By medical condition** | | | | | |
| HIV | 1,106,182 | 410 | 44,192 | 3.99 | 37.1 |
| ESRD | 803,055 | 254 | 46,595 | 5.80 | 31.6 |
| Diabetes | 31,733,668 | 2,251 | 2,227,132 | 7.02 | 7.1 |

Population sizes from: U.S. Census Bureau American Community Survey (nativity), CDC AtlasPlus (HIV), USRDS (ESRD), U.S. Diabetes Surveillance System (diabetes). TB cases from NTSS 2023. *Mtb* infection estimates from back-calculation, averaged over 2018–2021 to smooth COVID-19–related disruptions in TB surveillance during 2020–2021. *Mtb* prevalence = *Mtb* infections / population size $\times$ 100.

**Appendix Table 2.** Number needed to vaccinate to prevent one tuberculosis case by risk group, across vaccine efficacy assumptions of 40%–80%, in U.S. populations*

| Risk Group | VE = 40% | VE = 50% | VE = 60% | VE = 70% | VE = 80% |
|---|---|---|---|---|---|
| *All-comers vaccination* | | | | | |
| Total U.S. population | 87,409 (85,700–89,200) | 69,928 (68,600–71,300) | 58,273 (57,100–59,500) | 49,948 (49,000–51,000) | 43,705 (42,800–44,600) |
| Non-U.S.-born | 16,383 (16,000–16,800) | 13,106 (12,800–13,400) | 10,922 (10,700–11,200) | 9,362 (9,200–9,600) | 8,191 (8,000–8,400) |
| U.S.-born | 313,136 (301,000–326,000) | 250,509 (241,000–261,000) | 208,758 (201,000–218,000) | 178,935 (172,000–187,000) | 156,568 (150,000–163,000) |
| HIV | 6,745 (6,100–7,500) | 5,396 (4,900–6,000) | 4,497 (4,100–5,000) | 3,854 (3,500–4,300) | 3,373 (3,100–3,700) |
| ESRD | 7,904 (7,000–9,000) | 6,323 (5,600–7,200) | 5,269 (4,700–6,000) | 4,517 (4,000–5,100) | 3,952 (3,500–4,500) |
| Diabetes | 35,244 (33,800–36,700) | 28,195 (27,100–29,400) | 23,496 (22,600–24,500) | 20,139 (19,300–21,000) | 17,622 (16,900–18,400) |
| *Targeted vaccination* | | | | | |
| Total U.S. population | 2,783 (1,800–4,200) | 2,226 (1,400–3,300) | 1,855 (1,200–2,800) | 1,590 (1,000–2,400) | 1,391 (880–2,100) |
| Non-U.S.-born | 2,681 (2,100–3,400) | 2,145 (1,700–2,700) | 1,788 (1,400–2,300) | 1,532 (1,200–1,900) | 1,341 (1,000–1,700) |
| U.S.-born | 3,088 (950–7,600) | 2,471 (750–6,100) | 2,059 (630–5,100) | 1,765 (540–4,300) | 1,544 (470–3,800) |
| HIV | 269 (150–450) | 216 (120–360) | 180 (100–300) | 154 (86–260) | 135 (75–220) |
| ESRD | 459 (200–910) | 367 (160–730) | 306 (130–610) | 262 (110–520) | 229 (100–460) |
| Diabetes | 2,473 (1,600–3,700) | 1,979 (1,200–3,000) | 1,649 (1,000–2,500) | 1,413 (890–2,100) | 1,237 (780–1,900) |

*NNV, number needed to vaccinate; VE, vaccine efficacy; ESRD, end-stage renal disease; *Mtb*, *Mycobacterium tuberculosis.* Cells display NNV point estimate (95% CI). NNV estimates were generated by holding vaccine efficacy fixed at the indicated value while sampling TB case counts and the *Mtb*-infected subpopulation size from their respective bootstrap distributions (1,000,000 iterations). All-comers vaccination assumes vaccination of all individuals in the risk group regardless of *Mtb* infection status; targeted vaccination restricts vaccination to those with confirmed *Mtb* infection.

**Appendix Table 3.** Number needed to vaccinate (NNV) to prevent one outcome of interest, comparison of an M72/AS01$_E$-like tuberculosis vaccine to other established vaccines*

| **Vaccine** | **Outcome** | **Population** | **NNV (95% CI)** | **Reference** |
|---|---|---|---|---|
| *Tuberculosis (this study, targeted vaccination)* | | | | |
| M72/AS01$_E$ | TB disease prevention | PLWH, *Mtb*-infected | 217 (95–755) | This study |
| M72/AS01$_E$ | TB disease prevention | ESRD, *Mtb*-infected | 369 (133–1,370) | This study |
| M72/AS01$_E$ | TB disease prevention | Diabetes, *Mtb*-infected | 1,991 (960–6,800) | This study |
| M72/AS01$_E$ | TB disease prevention | Non-U.S.-born, *Mtb*-infected | 2,158 (1,200–7,100) | This study |
| M72/AS01$_E$ | TB disease prevention | U.S.-born, *Mtb*-infected | 2,486 (660–10,100) | This study |
| *Tuberculosis (this study, all-comers vaccination)* | | | | |
| M72/AS01$_E$ | TB disease prevention | PLWH, all | 5,429 (3,200–17,800) | This study |
| M72/AS01$_E$ | TB disease prevention | ESRD, all | 6,361 (3,700–20,800) | This study |
| M72/AS01$_E$ | TB disease prevention | Non-U.S.-born, all | 13,185 (7,800–42,700) | This study |
| M72/AS01$_E$ | TB disease prevention | U.S.-born, all | 252,021 (148,500–824,000) | This study |
| *Tuberculosis (modeled, Harris et al. 2020, China)* | | | | |
| TB vaccine (modeled) | TB disease prevention | 60–64y, pre-infection only | 1,022 | (*11*) |
| TB vaccine (modeled) | TB disease prevention | 60–64y, post-infection latency | 576 | (*11*) |
| TB vaccine (modeled) | TB disease prevention | 60–64y, post-infection L&R | 281 | (*11*) |
| TB vaccine (modeled) | TB disease prevention | 60–64y, pre- & post-infection | 230 | (*11*) |
| *Established adult vaccines* | | | | |
| Shingrix | Herpes zoster | Adults ≥50y | 12 (10–14) | (*12*) |
| Influenza | Influenza illness | Adults ≥65y (Australia) | 40 (15–250) | (*13*) |
| COVID-19 booster | Hospitalization | Adults | 205 (44–615) | (*14*) |
| PCV-13 | All CAP | Adults ≥65y (5y) | 234 | (*15*) |
| Zostavax | HZ hospitalization | Adults ≥65y | 263 (210–340) | (*16*) |
| PCV-13 | Hospital CAP | Adults ≥65y (5y) | 537 | (*15*) |
| HPV | Cervical cancer mortality | Girls vaccinated at 12y | 324 | (*17*) |
| HPV | Cervical cancer death | Adult women | 835 | (*17*) |
| Influenza | Hospitalization | Adults ≥65y | 850 (520–1,800) | (*18*) |
| PCV-13 | Outpatient CAP | Adults ≥65y (1y) | 1,287 | (*15*) |
| PCV-13 | Hospital CAP | Adults ≥65y (1y) | 1,620 | (*15*) |
| Influenza | Death | Adults ≥65y (Australia) | 5,000 (1,400–20,000) | (*13*) |

*NNV, number needed to vaccinate; CI, confidence interval; PLWH, persons living with HIV; ESRD, end-stage renal disease; *Mtb*, *Mycobacterium tuberculosis*; PCV-13, 13-valent pneumococcal conjugate vaccine; HPV, human papillomavirus; HZ, herpes zoster; CAP, community-acquired pneumonia; L&R, latent and recovered. NNV estimates from this study assume 50% vaccine efficacy and a 3-year time horizon. Comparator NNV values reflect universal or age-targeted vaccination programs and are shown for context; direct comparison with risk-targeted estimates from this study is limited by differences in target population and study design.

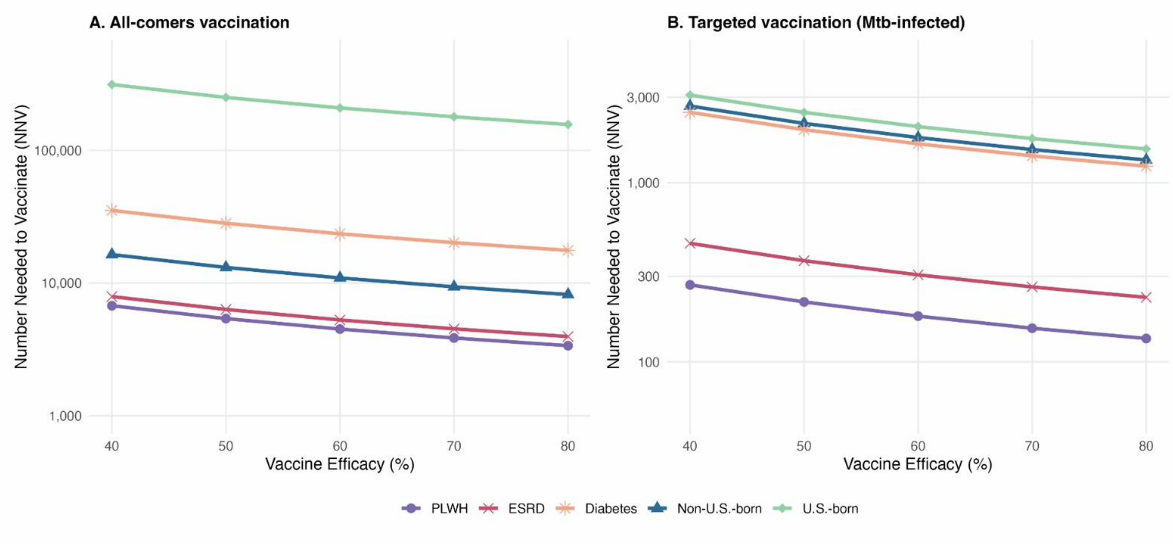


**Appendix Figure 1.** Number needed to vaccinate (NNV) by risk group across vaccine efficacy assumptions of 40%, 50%, 60%, 70%, and 80% in U.S. populations. Panel A shows NNV under all-comers vaccination (vaccinating all individuals in a risk group regardless of *Mtb* infection status); Panel B shows NNV under targeted vaccination (vaccinating only those with confirmed *Mtb* infection). NNV is plotted on a log scale. PLWH, persons living with HIV; ESRD, end-stage renal disease.
